# Changes in mobility pre and post first SARS-CoV-2 vaccination: findings from a prospective community cohort study including GPS movement tracking in England and Wales (Virus Watch)

**DOI:** 10.1101/2021.06.21.21259237

**Authors:** Vincent Nguyen, Yunzhe Liu, Richard Mumford, Ben Flanagan, Parth Patel, Isobel Braithwaite, Madhumita Shrotri, Thomas Byrne, Sarah Beale, Anna Aryee, Wing Lam Erica Fong, Ellen Fragaszy, Cyril Geismar, Annalan M D Navaratnam, Pia Hardelid, Jana Kovar, Addy Pope, Tao Cheng, Andrew Hayward, Robert W Aldridge

**Affiliations:** Centre for Public Health Data Science, Institute of Health Informatics, University College London, UK; Institute of Epidemiology and Health Care, University College London, London, UK; SpaceTimeLab, Department of Civil, Environmental and Geomatic Engineering, University College London, London, UK; Esri UK; Department of Infectious Disease Epidemiology, London School of Hygiene and Tropical Medicine, Keppel Street, London, UK; Department of Population, Policy and Practice, UCL Great Ormond Street Institute of Child Health, London, UK; Institute for Global Health, University College London, London, UK; Royal Free London NHS Foundation Trust, London, UK; Centre for Behaviour Change, University College London, London, UK; Francis Crick Institute, London, UK; Health Protection and Influenza Research Group, Division of Epidemiology and Public Health, University of Nottingham School of Medicine, Nottingham, United Kingdom; University College London Hospital, London, United Kingdom; Department of Computer Science, University College London, London, UK; London Centre for Nanotechnology and Division of Medicine, London, UCL

## Abstract

**Background:** Some evidence suggests that individuals may change adherence to public health policies aimed at reducing contact, transmission and spread of the SARS-CoV-2 virus after they receive their first SARS-CoV-2 vaccination. In this study, we aim to estimate the rate of change in average daily travel distance from a participant’s registered address before and after SARS-CoV-2 vaccination.

**Method:** Participants were recruited into Virus Watch starting in June 2020. Weekly surveys were sent out to participants and vaccination status was collected from January 2021 onwards. Between September 2020 and February 2021, we invited 13,120 adult Virus Watch participants to contribute towards our tracker sub-cohort which uses the Global Positioning System (GPS) to collect data on movement. We used segmented linear regression to estimate the median daily travel distance before and after the first self-reported SARS-CoV-2 vaccine dose.

**Results:** We analysed the daily travel distance of 228 vaccinated adults. Between 157 days prior to vaccination until the day before vaccination, the median daily travel distance travelled was 8.9km (IQR: 3.50km, 24.17km). Between the day of vaccination and 100 days after vaccination, the median daily travel distance travelled was 10.30km (IQR: 4.11, 27.53km). Between 157 days prior to vaccination and the vaccination date, there was a daily median decrease in mobility of 40m (95%CI: −51m, −31m, p-value <0.001) per day. After the removal of outlier data, and between the vaccination date and 99 days after vaccination, there was a median daily increase in movement of 45.0m (95%CI: 25m, 65m, p-value = <0.001). Restricting the analysis to the 3rd national lockdown (4th of January 2021 to the 5th of April 2021), we found a median daily movement increase of 9m (95%CI: −25m, 45m, p = 0.57) in the 30 days prior to vaccination and the vaccination date, and a median daily movement increase of 10m (95%CI: −60m, 94m, p-value = 0.69) in the 30 days after vaccination.

**Conclusions:** Our study demonstrates the feasibility of collecting high volume geolocation data as part of research projects, and the utility of these for understanding public health issues. Our results are consistent with both an increase and decrease in movement after vaccination and suggest that, amongst Virus Watch participants, any changes in movement distances post-vaccination are small.

## Introduction

The UK response to the COVID-19 pandemic has included restrictions on non-essential movement in order to reduce contacts and control transmission (1). However, the restriction of movement can have a detrimental impact on a wide variety of outcomes such as mental health, domestic accidents, the economy and education (2).

Vaccination against SARS-CoV-2 reduces COVID-19 transmission and disease (3) and as a result is a critical part of the strategies to allow more normal societal mixing. However, in the UK context, there are current concerns that misunderstandings about the effectiveness of the COVID-19 vaccine after the first dose may be leading to a reduction in adherence to other public health policies and increased exposure of partially protected individuals (4).

Preliminary research on vaccination in February 2021 found that 41% of over 80s who had their first dose of the vaccine had met another person within 3 weeks. These individuals did not include another household member, care worker or member of their support bubble, indoors which was permitted by the restrictions in place at the time(4). This is concerning as antibody levels will not have risen in the 1-2 weeks following the first dose of vaccine(5,6). Further evidence also suggests that those over the age of 80 are more likely to have a positive polymerase chain reaction test in the first 9 days of a vaccination compared to a control group, which might be explained by increased mobility and contacts between people in the period following vaccination(7).

With the emergence of the Delta (B.1.617.2) SARS-CoV2 variant, which is currently the dominant in the UK (8), the effectiveness of both the Oxford-AstraZeneca and Pfizer-BioNTech vaccines are estimated to be 33% against symptomatic disease after a first dose (9) although protection against hospitalisation appears to be much higher (10). Therefore, if those who are not fully vaccinated increase their level of social contact and mobility after the first dose, their risk of becoming infected and infecting others may also be increased.

Understanding movement post-first vaccination is important as it could help policy makers understand how perceived protection from the vaccination programme may negatively offset the effectiveness of other policies designed to reduce transmission. Whilst previous studies have attempted to investigate travel distances after vaccination(11), these were conducted using mobile call data based upon cellular tower location which is considered less accurate compared to GPS location.

In this analysis, we aim to quantify the effect that the first SARS-CoV-2 vaccination dose has on travelling behaviour using mobile phone global position system data collected from study participants that consented and voluntarily downloaded ArcGIS Tracker App onto their mobile phones.

## Methods

### Study Design

We conducted a quasi-experimental interrupted time series study.

### Data Resource

Virus Watch is a household community cohort study which began recruitment in June 2020 and has recruited 50,000 individuals by May 2021 across England and Wales with weekly online follow up, serology sampling and movement tracking in a subset of participants. A full description of the Virus Watch study has been published previously (12). Between September 2020 and February 2021, we invited 13,120 adults (aged 18+ on entry) from the Virus Watch study to contribute location data (longitude, latitude, date, time, travel mode and GPS accuracy which was defined by phone manufacturer’s GPS algorithm) using a proprietary mobile phone application (ArcGis Tracker).

### Intervention

Date of receiving the first dose of vaccine was self-reported through the weekly Virus Watch questionnaire. We began collecting weekly vaccination status on 11 January 2021 and asked about any prior vaccination during the first two weekly surveys. Subsequently, participants were asked to provide a weekly update only. The options available were “Pfizer/BioNTech”, “Oxford/AstraZeneca”, “Moderna” and “Other/Can’t remember”.

### Study Population

The study population included adults (18+ on entry) in the sub-cohort of the Virus Watch study who were vaccinated and submitted at least 10 days of readings. Participants had to submit 5 days of readings before and after their self reported vaccination date. We excluded readings which were outside of England and excluded analysis from days where there were less than 5 contributors. We only used location readings with an accuracy rating of less than 30 metres.

### Study Period

We started sending out invitations for the Tracker cohort between September 2020 and February 2021 with the data extract for this analysis being undertaken in May 2021.

### Outcomes

The primary outcome of the analysis was the change in median daily travel distance from a participant’s registered address. We choose the median daily distance to account for the distribution of the cohort’s daily travel patterns.

### Analysis

For each individual and each day, we calculated the cumulative outdoor travel distance recorded using the ArcGIS tracker app from their registered household address. This was calculated by summing up the distance (*d*) computed by the Euclidean distance method (*equation 1*) between the two sequential outdoor GPS records. Considering the accuracy of the GPS records, we set up a 25m radius buffer zone (the average horizontal accuracy is 25m) around a participant’s home location. Those points that fall within the buffer are considered as at-home travel activities and therefore considered as zero distance in analyses.

We defined the interruption time point in our analysis as the date of the first vaccination for each individual with negative days denoting dates before vaccination and positive days denoting days after vaccination. For each timepoint (days since vaccination), we calculated the median travel distance.

To calculate the trends in travel patterns, we used a segmented linear regression model; the first segment calculated travel trends before vaccination, and the second segment calculated trends after vaccination. To calculate the travel trajectory before vaccination, we conducted linear regression analysis to estimate the population’s median daily travel distance from home with time (days before vaccination) as the explanatory variable. To calculate the travel trajectory after vaccination, we conducted linear regression analysis using data after vaccination to estimate the population’s median daily travel distance from their home with time (days after vaccination) as the explanatory variable.

The UK vaccination programme prioritised people by (older) age and clinical risk groups, which, in addition to differences in the socio-economic backgrounds between those invited and accepting a vaccination, meant that selecting an appropriate control group for this analysis was not feasible.

### Covariates

Due to the study design, which compared the same individuals’ movement before and after vaccination, we did not use covariates for regression adjustment. For each eligible individual, we used the following data: days since vaccination and the total travel distance for the corresponding day.

### Sensitivity analyses

We performed two sensitivity analyses. First, after reviewing the data, we repeated the analyses with outliers removed. Second, to account for the effect of the removal of national restrictions on movement as alternative explanations for differences in movement after vaccination, we conducted a sensitivity analysis which limited travel and vaccination events to the third national lockdown. This period was between the 4th of January 2021 to the 5th of April 2021 and represents a time period when restrictions did not change in relation to rules about travel and social distancing.

### Ethics and Approval

The Virus Watch study was approved by the Hampstead NHS Health Research Authority Ethics Committee: 20/HRA/2320. All members of participating households provided informed consent for themselves and, where relevant, for children that they were responsible for. To contribute to the tracker subcohort of Virus Watch, adults had to provide explicit consent during our registration process.

### Information Governance

This research was registered with the UCL data protection office and reviewed by the UCL information security and governance teams. The Virus Watch Data Privacy Impact Assessment can be found online - https://ucl-virus-watch.net/?page_id=1228. During the consent and registration process, adult participants were invited to contribute geolocation data using the ArcGIS mobile phone tracker app. For those who chose to participate, we sent personal identifiable data from the UCL Data Safe Haven to a secure memory stick on a UCL machine from which we transferred the data via HTTPS into the ArcGIS Online subscription. The purpose of this data transfer was to set up participants’ tracker app accounts. The transferred data, along with the account passwords, were stored in North America. The UCL Virus Watch study team undertook the data transfer process and had access to the Participant Profile within the ArcGIS subscription. Only a small number of named ArcGIS employees had access to the participant profile area and only for the purposes of assisting the UCL study team when necessary. Once the tracker app accounts were created, the UCL study team emailed tracker app participants instructions on how to download the app and sign into the tracker app.

The geolocation data collected by the app was stored securely on a section of the ArcGIS Online Subscription hosted in Europe which is securely cleared every 30 days. Participants’ geolocation data was transferred on a regular basis via HTTPS to a secure memory stick on a UCL machine and was then imported via a secure gateway technology on to the UCL secure memory stick into the UCL Data Safe Haven.

Geolocation data was linked with other participant study data in the Data Safe Haven. Once analysed, aggregate data (generated from geolocation and other study data) was exported from the Data Safe Haven and published on the public study website and in research publications.

The aggregated dataset used in this analysis will be securely destroyed after 20 years in line with UCL’s record retention policy. In line with policies developed for electronic healthcare research, we did not report any data with a cell containing <5 events and where necessary, we protected these counts with secondary suppression.

## Results

79% of adult participants agreed to participate in the tracking subcohort of Virus Watch. We invited 13,120 participants into the Tracker sub-cohort and of these, 2,193 participants contributed at least 1 GPS reading. After removing invalid data points including those outside of England, and those which did not submit accurate readings (e.g., points exhibit extremely high horizontal and vertical accuracy), 1,376 participants were included. Of the 1,376 individuals, 1244 individuals were vaccinated by May 2021. After removing individuals with fewer than 5 data points either side of their vaccination date, 228 individuals were included in our final analysis.

Of the 228 participants, there were more females (57%) than males, with a median age of 62 (IQR: 55, 67). Individuals resident in local super output areas (LSOAs) in the three least deprived quintiles represented nearly 80% of the population. Nearly 90% of our cohort self-identified as “White - English/ Welsh/ Scottish/ Northern Irish/British”. See Table 1 for a socio-demographic breakdown of the cohort.

**Table 1:** Socio Demographic breakdown of the included cohort

| Characteristic | Sub Characteristic | N = 228 |
| --- | --- | --- |
| Gender at Birth | Male | 97 (43%) |
|  | Female | 131 (57%) |
| Median Age on Entry (IQR) |  | 62 (55, 67) |
| Index of Multiple Deprivation Quintile (LSOA of residence) | 1 (Most deprived) | 18 (7.9%) |
|  | 2 | 30 (13%) |
|  | 3 | 37 (16%) |
|  | 4 | 68 (30%) |
|  | 5 (Least deprived) | 75 (33%) |
| Region name | East Midlands | 33 (14%) |
|  | East of England | 44 (19%) |
|  | London | 36 (16%) |
|  | North East | 5 (2.2%) |
|  | North West | 22 (9.6%) |
|  | South East | 42 (18%) |
|  | South West | 18 (7.9%) |
|  | West Midlands | 16 (7.0%) |
|  | Yorkshire and The Humber | 12 (5.3%) |
| Ethnicity | White - English/Welsh/Scottish/Northern Irish/ British | 203 (89%) |
|  | Non - White British | 25 (11%) |

From the 228 participants, 157 days of eligible readings prior to the first vaccination and 100 days of eligible readings after the first vaccination were included. The mean number of people who contributed per day was 85.28 (95%CI: 78.78, 91.78). The mean number of days contributed by the 228 participants was 95.79 (95%CI: 90.28, 101.30).

Between 157 days prior to the first vaccination until the day before vaccination, the median daily travel distance was 8.9km (IQR: 3.50km, 24.17km). Between the day of the first vaccination until 100 days after that vaccination, the median daily travel distance was 10.30km (IQR: 4.11, 27.53km).

During the first segment of the linear regression model, from 157 days before the first vaccination until the vaccination date, there was a median daily decline of 40m (95%CI: −51m, −31m, p-value < 0.001) of movement per day (Figure 1a). During the second segment of the linear regression model, from the first vaccination date until 100 days after vaccination, there was a median daily increase of 114m (95%CI: 53.2m, 175.1m, p-value <0.001) in movement per-day.

**Figure 1:**
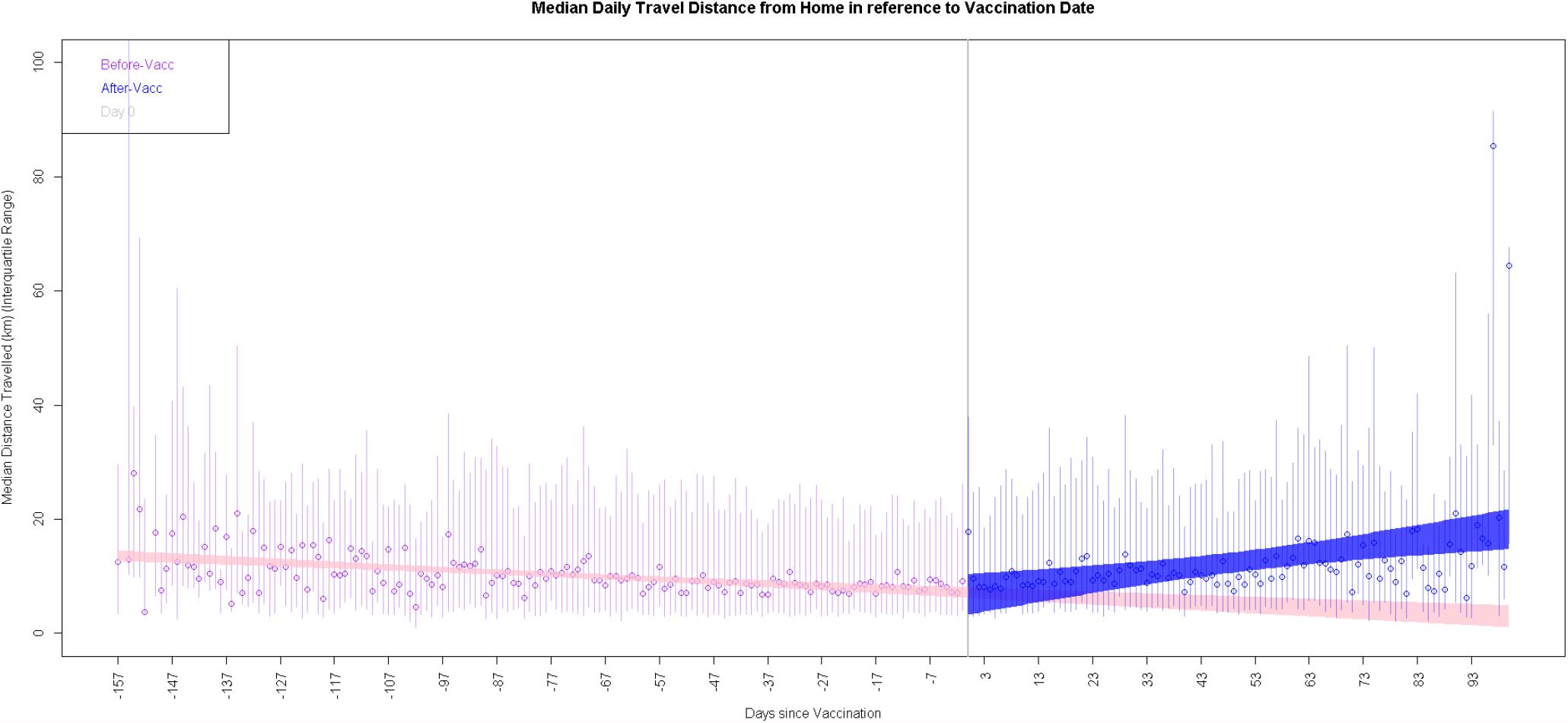
Graphical representation of the interrupted time series

**Figure 1b:**
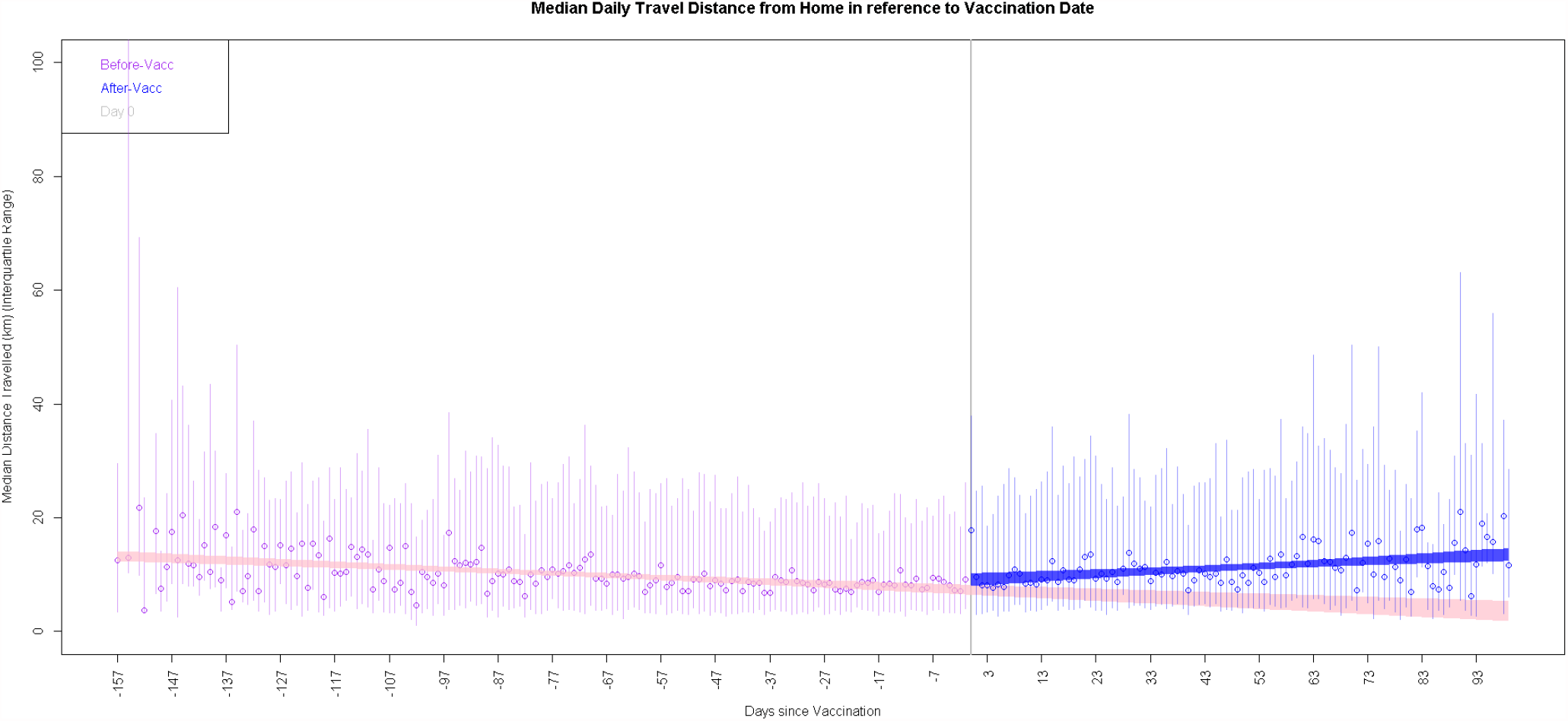
Graphical representation of the interrupted time series when outliers at the extreme end of the timeline

**Figure 1c:**
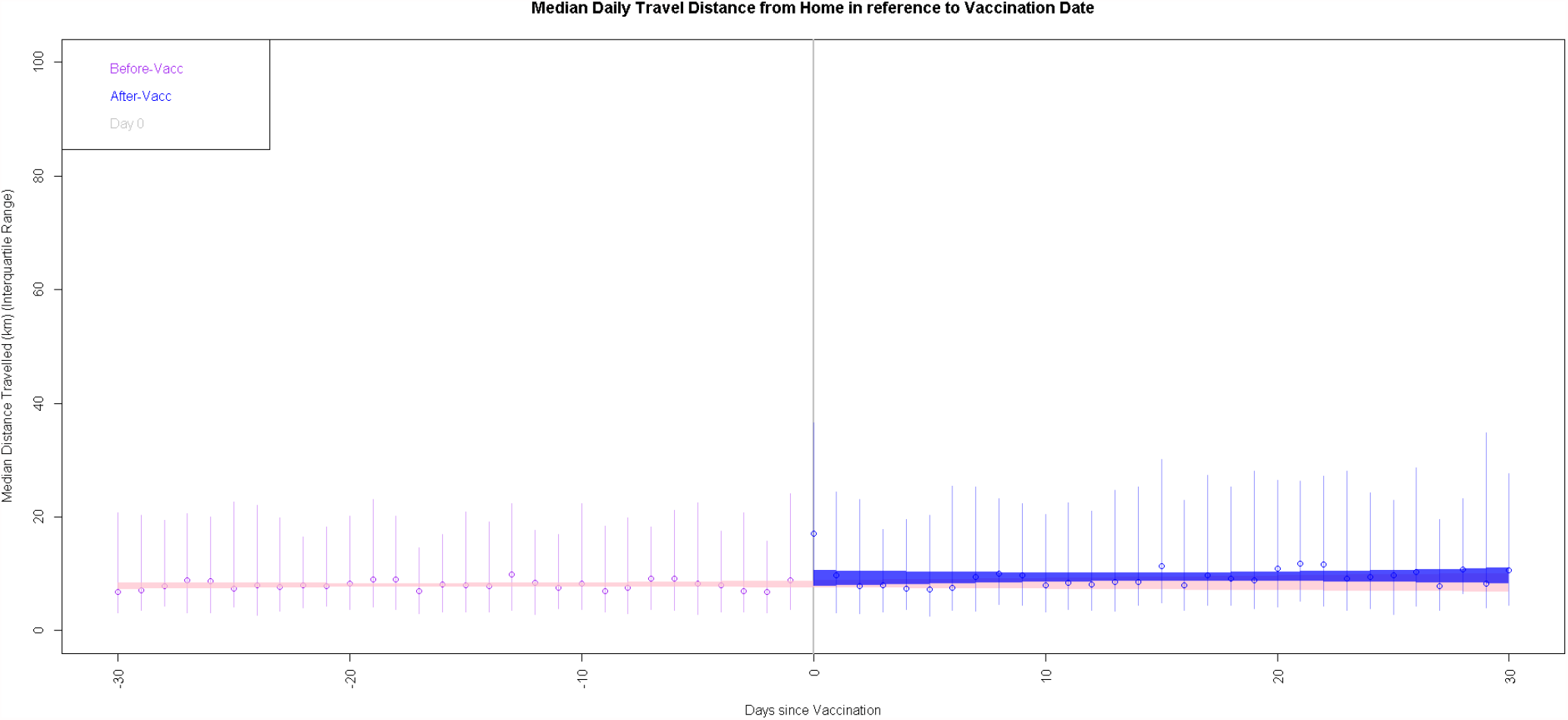
Graphical representation of the interrupted time series when only accounting for movement and vaccinations which occurred during the 3rd national lockdown (4th January 2021 - 5th April 2021) with a 30 day period either side of the vaccination date

### Sensitivity Analysis

After reviewing the data, we believe that certain values at either end of the timeline were outliers which we then removed. This changed the time interval to 157 days pre-vaccination with 99 days of followup post vaccination. In this sensitivity analysis, the median pre-vaccine travel distance between 157 before vaccination until the day before vaccination was 8.95km (IQR: 3.50km, 24.17km) and the median post-vaccine travel distance was 10.29km (IQR: 4.11km, 27.53km). Removing outliers updated the pre-vaccination segment to a daily decline of 37m (95%CI: −47.5m, −27.9m, p-value <0.001) and the post-vaccination segment to a daily increase of 45.1m (95%CI: 25.0m, 65.1m, p-value < 0.001). See figure 1b for a graphical representation of the interrupted time series when removing outlier values.

### Sensitivity Analysis 2

To account for the effect of national restrictions on movement, we conducted a sensitivity analysis which limited travel and vaccination events to the third national lockdown between 4th of January 2021 to the 5th of April 2021. Due to the asymmetry of the number of datapoints before and after vaccination in this analysis, we analysed movement 30 days before vaccination and 30 days after vaccination.

This reduced our cohort from 228 participants to 199 participants. Between 30 days prior to vaccination and the vaccination date, there was a median daily movement of 7.90km (95%CI: 3.43km, 19.94km) with a median daily movement increase of 9m (95%CI: −25m, 45m, p = 0.57).

Between the vaccination date and the following 30 days (during the 3rd national lockdown) there was a median daily movement of 9.12km (95%CI: 3.88km, 24.79km) with a median daily movement increase of 10m (95%CI: −60m, 94m, p-value = 0.69). See Figure 1c for a graphical representation of the interrupted time series for this sensitivity analysis.

## Discussion

Our study demonstrates the feasibility of collecting high volume geolocation data as part of research projects, and the utility of these for understanding public health issues. Our results require cautious interpretation. Our initial analysis found evidence of a modest increase in the rate of change in median daily distance travelled after participants received their first dose of SARS-CoV-2 vaccine, but when restricting our analysis to periods of lockdown, we did not find evidence of a difference in mobility following one vaccination dose. On balance, our results do not provide evidence that people increase the rate of their movements following first dose vaccination as they are consistent with both an increase and decrease in movement after vaccination and suggest that any change in mobility post vaccination is likely to be modest.

We used GPS data to measure the travel distance of vaccinated individuals. Not only does this improve accuracy over other methods of distance estimation by using the GPS system (as compared to cellular location), it also reduces recall bias when compared with using self reported data. Our interrupted time series study design controls for non time varying confounders as the same individual’s data are considered before and after vaccination. Our studies’ sample size means that we may be underpowered to detect small changes in mobility, particularly when restricting to the period of national lockdown. Our GPS collection was automated, but could be switched on and off by participants and is more likely to have been switched off on days when participants stayed at home. The use of the app in this way would result in our analysis over estimating the median distance travelled per day, through the non-reporting of GPS data (e.g. switching the app off) on days when participants stayed at home. Due to technological requirements of tracking applications, results were skewed towards those who had access to a smartphone and were able to contribute their data plan towards research activities, leading to low initial uptake rate. With a draining effect on battery life from the GPS app used, the drop-out rate from the tracker cohort of Virus Watch was relatively high. People taking part in Virus Watch are self-selecting and motivated to contribute to COVID-19 research and therefore their movement patterns may not be generalisable to all vaccinated groups.

Changes in national physical distance rules are likely to have led to an increase in the rate of change in median daily travel distance. It is also possible that other time varying confounders such as weather changes may affect the findings, with participants increasing mobility during this same time period post-vaccination as a result of improvements in weather. Whilst previous research found that nearly half of those who were vaccinated (after one dose) met with others outside their households or support bubbles (4), our findings provide a mixed picture about movement after the first dose of vaccination and we find no evidence of an increase when we conduct our analyses during periods of national movement restrictions.

Given previous studies have suggested people increase their non-household contacts after their first vaccination dose, further research on behaviour change following vaccination is warranted. In the meantime, it is important that public health communications are clear about the differential protection against SARS-CoV-2 infection offered by the first and second doses of the vaccine, such that people can exercise sound personal judgement on how they alter behaviour change following vaccination.

## Data Availability

We aim to share aggregate data from this project on our website and via a “Findings so far” section on our website - https://ucl-virus-watch.net/. We will also be sharing individual record level data with personal identifiers removed on a research data sharing service such as the Office of National Statistics Secure Research Service. In sharing the data we will work within the principles set out in the UKRI Guidance on best practice in the management of research data. Access to use of the data whilst research is being conducted will be managed by the Chief Investigators (ACH and RWA) in accordance with the principles set out in the UKRI guidance on best practice in the management of research data. It is the intention that the data arising from this research will initially be collected, cleaned and validated by the UCL research team and once this has been completed will be shared for wider use. We aim to make subsets of the data more rapidly available both on our study website and via the public facing dashboard during the ongoing phase of data collection. In line with Principle 5 of the UKRI guidance on best practice in the management of research data, we plan to release data in batches as they become available or as updated results are published. Individual record data linked using NHS Digital will not be shared, only aggregated results. HES and mortality data may be obtained from a third party and are not publicly available. These data are owned by a third party and can be accessed by researchers applying to the Health and Social Care Information Centre for England. We will put analysis code on publicly available repositories to enable their reuse.

## Tables, Equations and Graphs

Equation 1: Equation to calculate the distance travelled by each participant

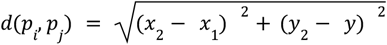

Where *d(p*_*i*_,*p*_*j*_ *) is the Euclidean distance between two sequential GPS points (i*.*e*., *p*_*i*_ *and p_j_); the Cartesian coordinates of p*_*i*_ *are (x*_*1*_, *y*_*1*_) *and (x*_*2*_, *y*_*2*_*) for p*_*j*_. *We employed the British National Grid (BNG) as the reference system*.

